# Postmortem Validation of Quantitative MRI for White Matter Hyperintensities in Alzheimer’s Disease

**DOI:** 10.1101/2025.08.05.25332820

**Authors:** Mariam Mojtabai, Rajeswar Kumar, Nicolas Honnorat, Karl Li, David H. Wang, Jinqi Li, Ray F. Lee, Timothy E. Richardson, José E. Cavazos, Mustapha Bouhrara, Jon B. Toledo, Susan Heckbert, Margaret E. Flanagan, Kevin F. Bieniek, Jamie M. Walker, Sudha Seshadri, Mohamad Habes

## Abstract

White matter hyperintensities (WMH) are frequently observed on MRI in aging and Alzheimer’s disease (AD), yet their microstructural pathology remains poorly characterized. Conventional MRI sequences provide limited information to describe the tissue abnormalities underlying WMH, while histopathology—the gold standard—can only be applied postmortem. Quantitative MRI (qMRI) offers promising non-invasive alternatives to postmortem histopathology, but lacks histological validation of these metrics in AD. In this study, we examined the relationship between MRI metrics and histopathology in postmortem brain scans from eight donors with AD from the South Texas Alzheimer’s Disease Research Center. Regions of interest are delineated by aligning MRI-identified WMH in the brain donor scans with postmortem histological sections. Histopathological features, including myelin integrity, tissue vacuolation, and gliosis, are quantified within these regions using machine learning. We report the correlations between these histopathological measures and two qMRI metrics: T2 and absolute myelin water signal (aMWS) maps, as well as conventional T1w/T2w MRI. The results derived from aMWS and T2 mapping indicate a strong association between WMH, myelin loss, and increased tissue vacuolation. Bland-Altman analyses indicated that T2 mapping showed more consistent agreement with histopathology, whereas the derived aMWS demonstrated signs of systematic bias. T1w/T2w values exhibited weaker associations with histological alterations. Additionally, we observed distinct patterns of gliosis in periventricular and subcortical WMH. Our study presents one of the first histopathological validations of qMRI in AD, confirming that aMWS and T2 mapping are robust, non-invasive biomarkers that offer promising ways to monitor white matter pathology in neurodegenerative disorders.

## 1. Introduction

White matter hyperintensities (WMH), characterized by increased signal intensity on T2-weighted MRI scans (Habes, et al. 2016), are frequently observed in normative aging and Alzheimer’s disease (AD) and correlate with cognitive impairment and clinical progression (Prins and Scheltens 2015). The precise microstructural pathology underlying WMH in AD remains elusive despite its prevalence (Pradeep, et al. 2024, Gouw, et al. 2008, Solé-Guardia, et al. 2025). Traditional magnetic resonance imaging (MRI) methods, which rely primarily on qualitative assessments and direct signal evaluation, lack specificity in distinguishing critical tissue changes, such as myelin loss or gliosis, each of which carries distinct implications for disease mechanisms. This limitation is not unexpected, as conventional MRI signal intensity is inherently relative and lacks direct physical specificity (Bloem, et al. 2018).

Previous histopathology studies have shown associations with increased arteriolosclerosis (McAleese, Miah, et al. 2021), neurofibrillary tangle spread (Braak staging) (Braak and Braak 1991), myelin/axonal loss (Silbert, et al. 2024), and gliosis (Erten-Lyons, et al. 2013). Postmortem histopathological examination is the gold standard to characterize specific tissue pathology associated with WMH; however, this approach is invasive, labor-intensive, and can only be carried out after death (Jack Jr, et al. 2018). Consequently, there is an urgent need for validated, non-invasive in vivo imaging biomarkers capturing the tissue changes underlying WMH (Li, et al. 2023).

Quantitative MRI (qMRI) methods were developed to produce stable MRI metrics better reflecting the composition of biological tissues than standard radiological scans (Seiler, Nöth, et al. 2021). The signal intensities of radiological scans cannot be directly compared across scanners and can even vary between subjects using the same scanner (Ruder, et al. 2012). qMRI often involves an additional analysis step conducted on standard MRI data, generating new values that can be interpreted in a physically meaningful way. This approach includes T2 mapping (O’Brien, et al. 2022) and apparent myelin water signal (aMWS) mapping (Lee, et al. 2021). qMRI methods offer promising alternatives to postmortem histopathology by providing quantitative assessments sensitive to microstructural changes in brain tissues. They have demonstrated potential in other neurological conditions to detect myelin integrity (Piredda, et al. 2021), tissue water content (Abbas, et al. 2015), and microstructural damage (Seiler, Nöth, et al. 2021). Unfortunately, their validation through direct histological comparison, particularly in AD, remains scarce. This issue has limited their clinical translation and their use as biomarkers for the aging brain.

In this study, we address this concern by investigating the postmortem brain samples of eight participants clinically diagnosed with AD at the South Texas Alzheimer’s Disease Research Center (Li, et al. 2023). Each brain sample was scanned to acquire a comprehensive set of MRI images prior to dissection and histopathological analysis (Li, et al. 2023). We applied three different processing methods to the MRI images to derive two quantitative MRI (qMRI) measures: T2 mapping and aMWS mapping, and another commonly used, but nonquantitative method: T1-weighted/T2-weighted (T1w/T2w) MRI. We quantify the ability of these MRI measures to detect histopathological changes in WMH by comparing the MRI measures with histopathological features extracted via machine learning from the digitized optical microscopy images of six different histopathology stains. Overall, our study represents one of the first attempts to precisely validate the ability of MRI measures to track the neuropathological alterations underlying the WMH associated with AD.

## 2. Methods

### 2.1 Brain Samples

The neuropathology core at the UTHSCSA Biggs Institute has an active brain donation program (Li, et al. 2023). All brain samples are processed following the National Alzheimer’s Coordinating Center (NACC) protocols (Beekly, et al. 2007). Donors and/or legally authorized representatives have consented to the broad sharing of biospecimens and data. Following the Biggs Brain Bank protocol, the brain and dura undergo a detailed external examination. The brain is then bisected along the midsagittal plane and the left cerebral and cerebellar hemispheres. The right side of the brain is sectioned into 1 cm slabs, frozen between aluminum plates, and stored in a -80° C freezer at the Biggs Brain Bank for future studies. The left hemisphere is immersed in a 10% neutral buffered formalin solution and imaged in an MRI scanner belonging to the UTHSCSA Research Imaging Institute before undergoing dissection and neuropathological examination (Li, et al. 2023) (Figure 1).

**Figure 1:**
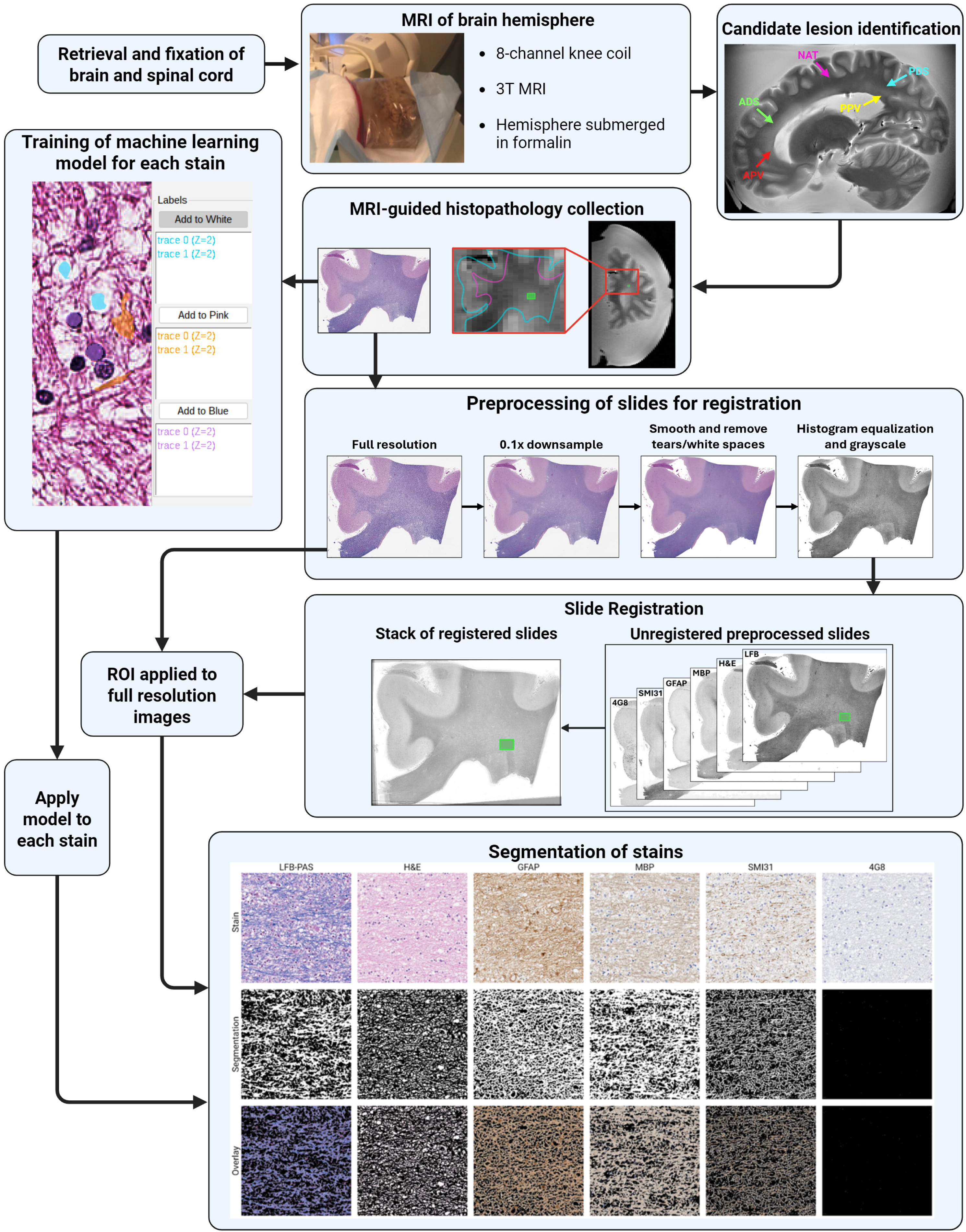
Study Overview. After formalin fixation, one hemisphere of each brain was scanned using MRI. These scans were used to identify WMH. The brain was then sectioned into coronal slabs, and the MRI data guided the localization of WMH on the slabs. Regions containing WMH were sampled by cutting out corresponding tissue sections. The selected histological sections were digitized and used to train machine learning models to segment and quantify specific features associated with each histological stain. To ensure consistent analysis across stains, the slides were registered to one another, allowing for accurate placement of the region of interest (ROI) as indicated by the MRI. Once the ROIs were established on the histological images, the trained models were applied to extract quantitative features. Segmentations for each target color were generated, and overlays of these segmentations on the original stains are provided.

Sample collection at the Brain Bank is ongoing (Li, et al. 2023). For this project, eight brain samples were selected for detailed histological examination. These brains were selected among brain samples presenting both anterior WMH (in the frontal lobe near the end of the anterior horn of the lateral ventricle) and posterior WMH (in the posteromedial aspect of the parietal lobe near the posterior horn of the lateral ventricle) as well as normal-appearing tissue near the base of the postcentral gyrus, which was considered as a control healthy tissue during our experiments. The formalin fixation times between autopsy, postmortem MRI scans, and histopathological dissection for these eight brain samples are reported in Table 1, along with donor demographics and their diagnoses.

**Table 1:** The eight brain samples considered in this study. Clinical diagnoses and diagnoses derived from the neuropathological examinations are listed for each case. Considering the autopsy as day 0, scanning and dissection times are reported in the last two columns. Brain samples were fixed in formalin during these time periods.

| Case | Sex | Age Range at Death | Clinical Diagnosis | Pathological Diagnosis | Scan Day | Dissection Day |
| --- | --- | --- | --- | --- | --- | --- |
| 1 | M | 81-85 | AD | AD, LATE-1, Severe VD | 44 | 114 |
| 2 | M | 76-80 | AD | AD, LBD-3, LATE-1, Moderate VD | 28 | 39 |
| 3 | F | 61-65 | AD | AD, Mild VD | 86 | 98 |
| 4 | M | 85-90 | AD | AD, LATE-1, Moderate VD | 76 | 138 |
| 5 | M | 61-65 | AD | AD, LATE-1, Moderate VD | 87 | 127 |
| 6 | F | 71-75 | AD | AD, LBD-2, LATE-2, Mild VD | 73 | 162 |
| 7 | F | 76-80 | VD, AD | AD, LATE-2, Moderate VD | 147 | 162 |
| 8 | M | 76-80 | AD | AD, Severe VD | 17 | 142 |

### 2.2 Image Acquisition

The scans were acquired using a Siemens TIM TRIO 3T scanner equipped with an 8-channel, phased-array volume coil (knee coil). The postmortem MRI acquisition protocol included a T1-weighted sequence with a repetition time (TR) of 2200 ms, an echo time (TE) of 3.25 ms, a flip angle of 15 degrees, and a resolution of 0.5 × 0.5 × 0.5 mm³. T2-weighted images were also acquired using a turbo spin echo (TSE) sequence, with a TR of 3750 ms, TEs of 20 ms, 30 ms, and 50 ms, a flip angle of 20 degrees, and a resolution of 1.5 × 1.0 × 1.0 mm³ (Li, et al. 2023). As mentioned above, the scans were performed on formalin-fixed individual hemispheres suspended in formalin. Between 2019 and 2025, more than 300 brain samples were imaged using this protocol at the Biggs Institute, including the eight samples considered in this work, before their dissection.

### 2.3 WMH Annotation and Selection

The T2-weighted scans corresponding to the shortest echo time (TE = 20 ms) were manually annotated by the neurologist in our group, to select four distinct WMH regions in each brain sample: anterior periventricular (APV), anterior deep subcortical (ADS), posterior periventricular (PPV), posterior deep subcortical (PDS), and normal-appearing tissue (NAT) regions. NAT regions were placed at the base of the postcentral gyrus to serve as controls. These T2-weighted scans were selected because we observed a stronger contrast between grey matter and white matter in these scans compared to the T2-weighted scans acquired for the longer echo times of 30 ms and 50 ms. T1-weighted MRI scans were consulted to confirm these selections and ensure that the regions did not contain brain lesions or ventricular spaces.

### 2.4 Quantitative MRI

The calculation of the qMRI measures was carried out separately for each brain sample by aligning the three T2-weighted MRI images and the T1-weighted image using the ANTsPy (version 0.4.2) rigid registration method (Avants, et al. 2011). Since the ROIs were placed in the T2-weighted image corresponding to the shortest echo time (TE = 20 ms), the T1-weighted and the remaining T2-weighted scans were registered in that space. The transformation calculated for the T1-weighted scan was applied to a brain mask manually delineated in the T1-weighted scan by an expert to mask out the formalin signal (Li, et al. 2023). This operation produced a warped mask aligned with the T2-weighted scan acquired with the shortest echo time. This warped mask was multiplied by the four aligned scans, and the qMRI measures were derived. To correct for intensity non-uniformity, the N4ITK (version 2.5.0; ITK version 5.4) bias field correction method (Tustison and Gee 2009) was applied to the average of the three T2-weighted echo images for each subject. The resulting bias field was then used to uniformly correct all three echo images for that subject. Two qMRI methods were investigated in this study: T2 mapping and aMWS mapping. In addition, a commonly used method to map myelin T1w/T2w MRI was explored.

#### 2.4.1 T2 Mapping

T2 mapping, a widely used qMRI technique, quantifies the transverse relaxation time (T2) in each MRI voxel and displays the quantitative T2 values as an intensity-based map (O’Brien, et al. 2022). In standard T2-weighted scans, the intensity value is a relative value that depends on scanner and sequence parameters. By contrast, T2 mapping produces T2 times reflecting the intrinsic properties of brain tissues, which can be compared between scans and scanners (O’Brien, et al. 2022). T2 maps are typically derived from spin-echo sequences (McRobbie, Moore, et al., MRI from Picture to Proton 2017) implementing multiple different echo times (TE). These different echo times produce images presenting different signal intensities. For each brain location and at first approximation, the signal intensity at different echo times is usually modeled using the following exponential decay function:

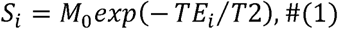

where *S_i_* represents the signal intensity at the *i*-th echo time, *M_0_* is the equilibrium magnetization, *TE_i_* is the *i*-*th* echo time, and T2 is the composite transverse relaxation time from all water compartments in the tissue (McRobbie, Moore, et al. 2017). Voxel-wise T2 values are calculated and mapped by fitting this model.

During our experiments, an average quantitative T2 value was calculated separately for each manually delineated WMH region, as illustrated in Figure 2 (row 1). These averages were compared with the histopathology features.

**Figure 2:**
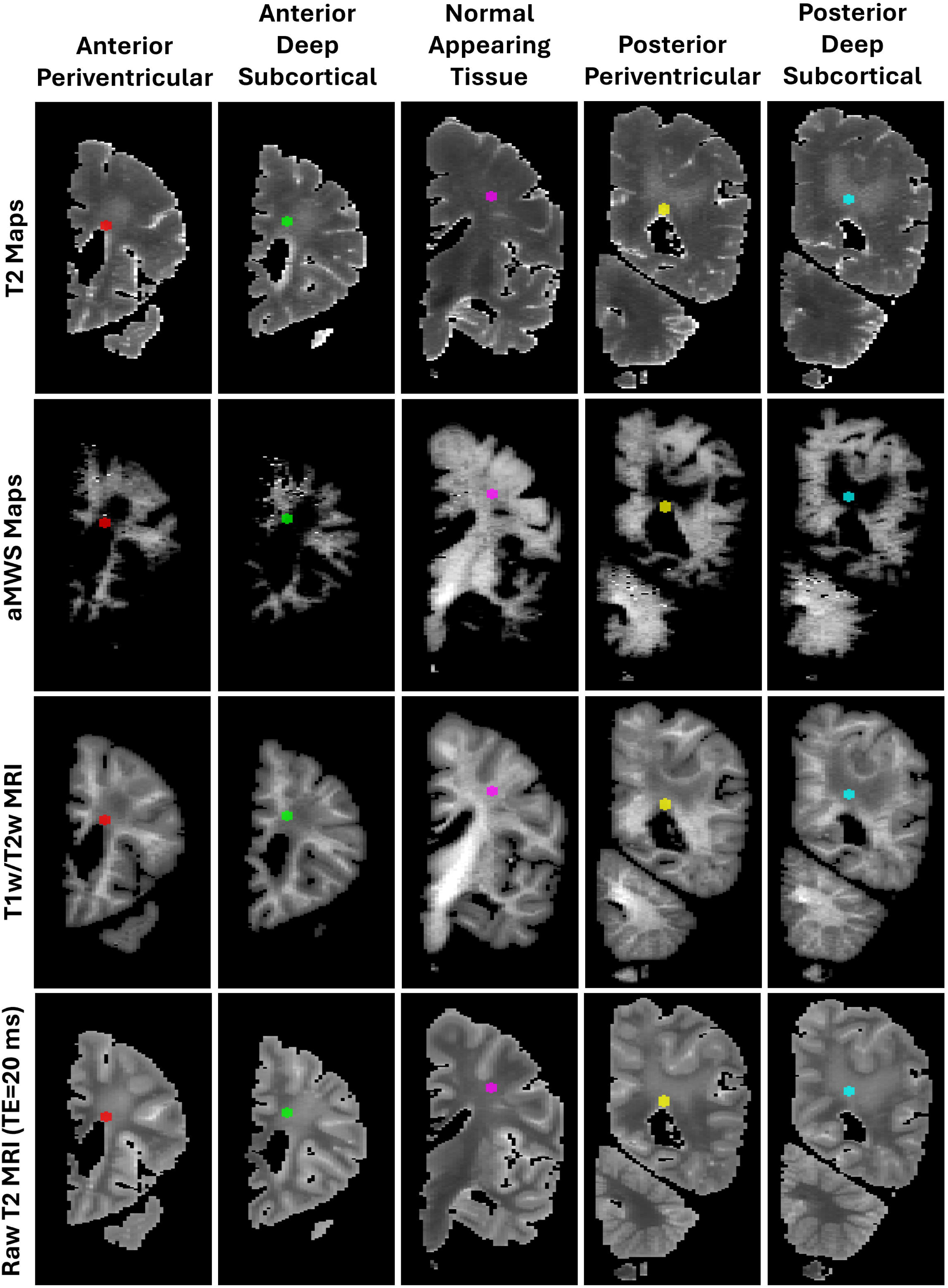
qMRI maps derived for the first brain sample and annotated with the five selected WMH regions: anterior periventricular (APV), anterior deep subcortical (ADS), normal-appearing tissue (NAT), posterior periventricular (PPV), and posterior deep subcortical (PDS). The NAT region was selected at the base of the postcentral gyrus.

#### 2.4.2 Apparent Myelin Water Signal (aMWS) Mapping

The aMWS maps generated were inspired by a quantitative MRI method used in previous studies (Lee, et al. 2021, MacKay and Laule 2016) that analyzes the myelin content of brain tissue. It relies on the fact that water trapped between myelin layers presents a significantly shorter T2 relaxation time compared to water in the extracellular and intracellular spaces. Like T2 mapping, this method models the decay of the MRI signal over time. However, instead of focusing solely on the T2 relaxation time, this technique isolates the signal contribution from myelin water. This method provides a measure of myelin water fraction (MWF) representing the ratio of the signal from myelin water to the total MRI signal, producing a voxel-wise map that reflects the relative myelin content within brain tissue. This method has emerged as a critical tool for measuring myelin integrity and studying brain disorders that induce significant damage to neurons’ myelin sheaths (Faulkner, et al. 2024).

Various models have been proposed in the literature (Alonso-Ortiz, Levesque and Pike 2015). Due to the constraint of having just three echo times, a modified version of the two-compartment model (West, et al. 2018) was used here to derive the aMWS maps. Two-compartment models assume that one compartment contains water trapped in myelin, while the water in the second compartment is not affected by the myelin (Does 2018, Whittall and MacKay 1989). Short T2 times are expected for the water in the myelin compartment, whereas longer T2 times should be observed in the second compartment. The MRI signal under the modified two-compartment model is modelled as follows:

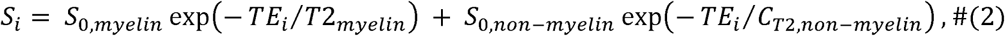

where *S_0,myelin_*and *S_0,non-myelin_* are the extrapolated signals at TE = 0 in the myelin and non-myelin water, respectively. T2_myelin_ is the T2 time for the compartment containing the myelin water. To ensure a well-posed fit with only three available echo times, the number of free parameters was limited to three. Specifically, *C_T2,non-myelin_* was fixed to a constant value. This value was estimated using gray matter and white matter masks, generated for each subject using a previously described method (Honnorat, et al. 2025). These masks were applied to the T2 maps, and the average T2 value within the gray matter was calculated, resulting in a *C_T2,non-myelin_* of 70 ms. Gray matter was chosen as the reference tissue because it contains minimal myelin, making its T2 relaxation time a reasonable approximation for the non-myelin water pool.

Once the signal from each compartment is determined, the apparent signal of myelin water is calculated using the derived signals for the myelin and non-myelin water (West, et al. 2018):

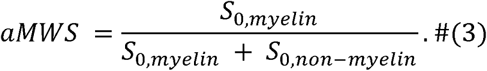

During our experiments, aMWS values were averaged within the manually delineated WMH regions, following the same approach used for the quantitative T2 maps. This approach is shown in Figure 2 (row 2).

### 2.5 T1w/T2w MRI

T1w/T2w MRI has been widely used as a method to estimate myelin density (Glasser and Van Essen 2011, Glasser, Goyal, et al. 2014). These myelin maps are created by aligning the T1-weighted and T2-weighted MRI scans and dividing the T1-weighted signal by the T2-weighted signal (TE = 20 ms) in each voxel. During our experiments, an average T1w/T2w ratio was calculated for each WMH region (Figure 2, row 3). These averages could be compared across brain samples because all our scans were acquired within the same scanner.

### 2.6 Histopathological Examination

#### 2.6.1 Histopathology Procedure

The first step of the dissection protocol implemented at the Biggs Brain Bank involves sectioning the brain samples into coronal slabs of 1 cm thickness (Li, et al. 2023). The brain slabs corresponding to the WMH regions delineated by our neurologist in the T2-weighted scans were identified by visually comparing anatomical landmarks easily noticeable in both the MRI scans and the physical brain, and gross anatomical landmarks (thalamus, postcentral sulcus, parieto-occipital sulcus). The exact WMH locations were selected with nearby macro- (gyri and ventricle shape) and micro-landmarks (enlarged perivascular spaces, lacunes, if available). For each selected slab, a small, squared section of around 2 to 3 cm was cut inside the WMH region, and six histological slides were prepared using six different stains.

These six stains were analyzed to understand the histological substrates of the WMH regions. Hematoxylin and eosin (H&E) stain the cell nucleus, extracellular matrix, and cytoplasm (Feldman and Wolfe 2014) to provide a broad overview of histological features. Luxol fast blue (LFB), while not specific, stains myelin (Klüver and Barrera 1953) to identify myelin pallor. The LFB stains were combined with periodic acid-Schiff (PAS) to visualize the tissue around the myelin. Myelin basic protein (MBP) antibody targets the MBP involved in myelin formation and maintenance (Sternberger, et al. 1978) while the SMI31 antibody targets phosphorylated neurofilament, a marker of axons (Rudrabhatla, Jaffe and Pant 2011). Combining their results informs whether the myelin pallor is due to just demyelination or if axonal loss is involved. Glial fibrillary acidic protein (GFAP) antibody stains astrocytes, identifying glial cells (Eng and Ghirnikar 1994) to identify areas of increased gliosis. Finally, the 4G8 antibody targets the Aβ protein (Taddei, et al. 2022), and allows the examination of any local amyloid angiopathy within the blood vessels. The stained slides obtained using these six stains were digitally scanned using a Leica Biosystems Aperio CS2 digital scanner with 20x magnification capabilities.

#### 2.6.2 Machine Learning Analysis

To quantitatively assess the extent of the stains, a random forest algorithm was trained using the Trainable Weka Segmentation plugin in ImageJ (Arganda-Carreras, et al. 2017) to segment each type of slide. The default training features were used, including Gaussian blur, hessian, membrane projections, Sobel filter, and Gaussian differences. A batch size of 100, 24 threads, and 20 trees was used when training the model. A sample of each stain was taken from each brain sample to train the models to ensure a diverse training set. ROIs were manually placed to cover most of the stain colors and texture variations when training the random forests. Following the initial training based on these manual segmentations, models’ outputs were reviewed, and segmentation errors were corrected. The models were retrained iteratively until they achieved a satisfactory accuracy in segmenting the stains.

In H&E staining, the hematoxylin stains the cell nuclei a blue/dark purple color, and the eosin stains the extracellular matrix and cytoplasm a pink/red color. The random forest was then trained to segment the pink, blue, and white pixels. The white pixels correspond to the space between tissues and were counted to measure vacuolation. LFB stains the myelin blue while the PAS stains the tissue a pink to dark purple color. A random forest model was then trained to detect these three colors and the white spaces between the tissue. The area of blue stain was used to measure the extent of myelin. MBP, GFAP, and SMI31 are immunohistochemical stains that utilize the binding of antigens to specific antibodies (Ramos-Vara 2005). The target antibody determines what will be stained. The appearance of these stains is brown. The machine learning models were then trained to segment brown and white spaces. Table 2 summarizes how changes in each stain are interpreted in the context of normal-appearing tissue and areas with WMH.

**Table 2:** Biological interpretation of the histopathology stains.

| Histopathology Stain | % Area Increase Indicates | % Area Decrease Indicates | Signal direction reflecting NAT |
| --- | --- | --- | --- |
| LFB (blue-stained tissues) | Preserved myelinated tissue | Demyelination, neurodegeneration | ↑ is better |
| H&E (unstained tissues) | Tissue vacuolation, rarefaction, or cell loss | Preserved cellular architecture | ↓ is better |
| GFAP (stained tissues) | Increased astrocytosis; gliosis response to injury | Reduced astrocytic activity; possibly normal tissue or severe damage with glial loss | ↓ is better |
| MBP (stained tissues) | Preserved myelin | Myelin loss or demyelination | ↑ is better |
| SMI31 (stained tissues) | Preserved axons | Axonal loss | ↑ is better |
| 4G8 (stained tissues) | Presence of vascular amyloid pathology | Low or absent amyloid pathology | ↓ is better |

Besides their automatic segmentation, the digital images of the six slides considered for each WMH region within each brain sample were also down-sampled and aligned as follows. The six slides were down-sampled by a factor of ten, smoothed using a Gaussian filter with a standard deviation (σ) of 5 pixels, converted to grayscale, and their histograms were equalized using the contrast-limited adaptive histogram equalization (CLAHE) method (Zuiderveld and others 1994) (Figure 1). This preprocessing enhanced the contrast between gray and white matter. Then, the Greedy non-rigid registration tool (Venet, et al. 2021) was used to align all the stains with the LFB stain, which usually exhibited the best contrast between white matter and gray matter.

The stack of aligned slides was manually reoriented to match the MRI, and this rough alignment was used to guide the manual selection of a region of interest within the stack of aligned stain slides. This ROI was saved as a binary mask, and the inverses of the registration transforms generated by Greedy were applied to project this binary mask over the original histology slides. Histopathological features were defined by measuring the proportion of the area of these projected binary masks that was covered by cell structures detected by the random forests. For instance, in the H&E histology slides, the proportion of binary mask pixels detected as background/white pixels by the random forest trained to segment the H&E slides was calculated and reported as a single feature, the *percentage of vacuolation*. Similarly, the percentage of the mask area covered by cell structures containing myelin was obtained by focusing on the blue structures revealed by the LFB stain. The interpretation of the other stains is presented in Table 2.

### 2.7 Statistical Analysis

Spearman correlation tests were used to assess the relationship between qMRI measures and histological features. More specifically, for each stain and each qMRI measure, a Spearman correlation coefficient and a p-value were calculated, for a statistical significance set at p < 0.05. To maximize statistical power, all the regions containing ROIs were included in the analysis (Figure 3). Bland–Altman plots were generated to assess the agreement between qMRI measures and histopathology features after z-scoring the data (Figure 4).

**Figure 3:**
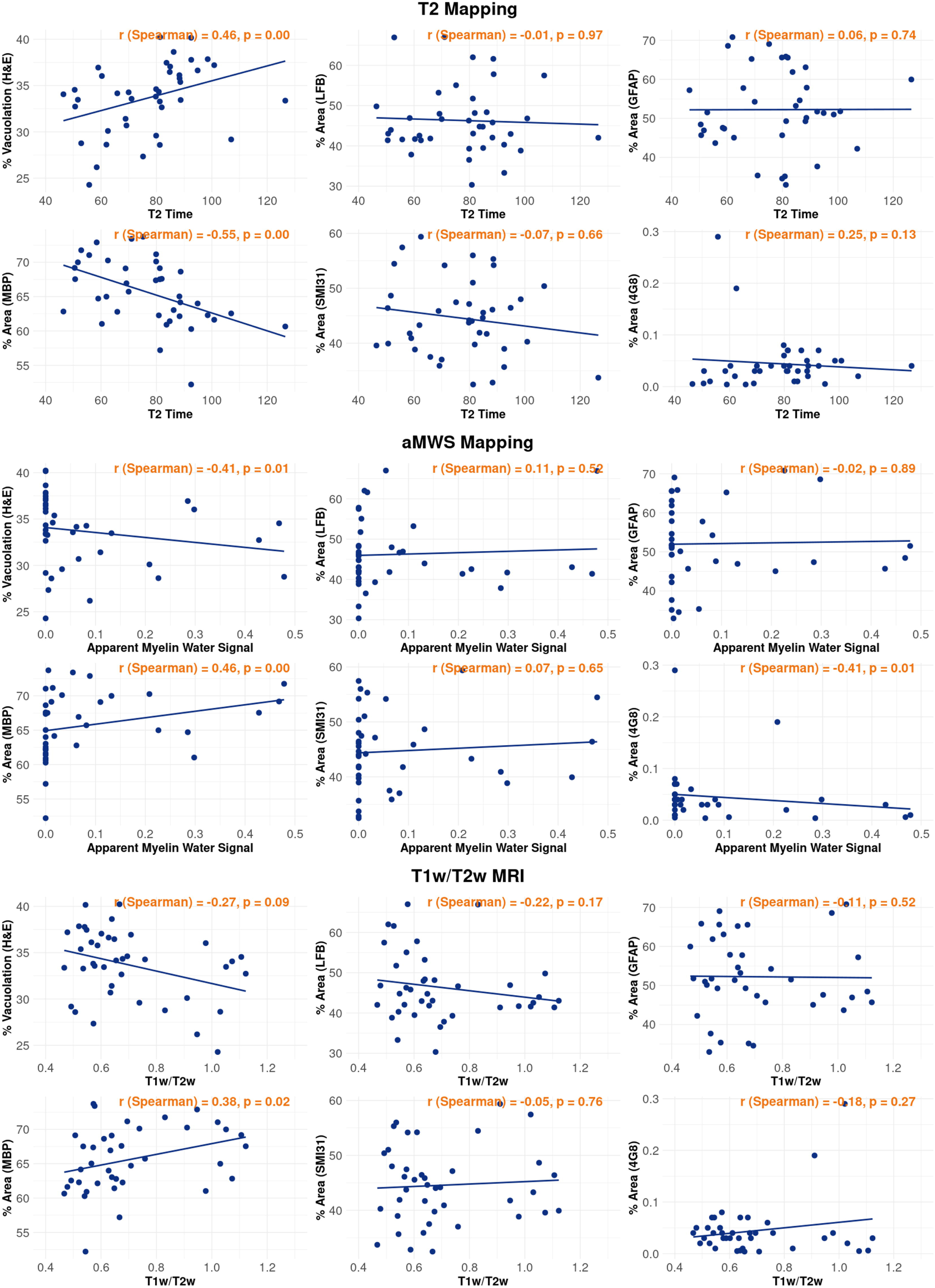
Correlations between histological features and qMRI measures (T2 mapping: first block, aMWS mapping: second block, T1w/T2w MRI: third block). The interpretation of each stain’s directionality is summarized in Table 2. Higher values of LFB, MBP, and SMI31 indicate better white matter integrity. Higher values of H&E, GFAP, and 4G8 indicate worse white matter integrity.

**Figure 4:**
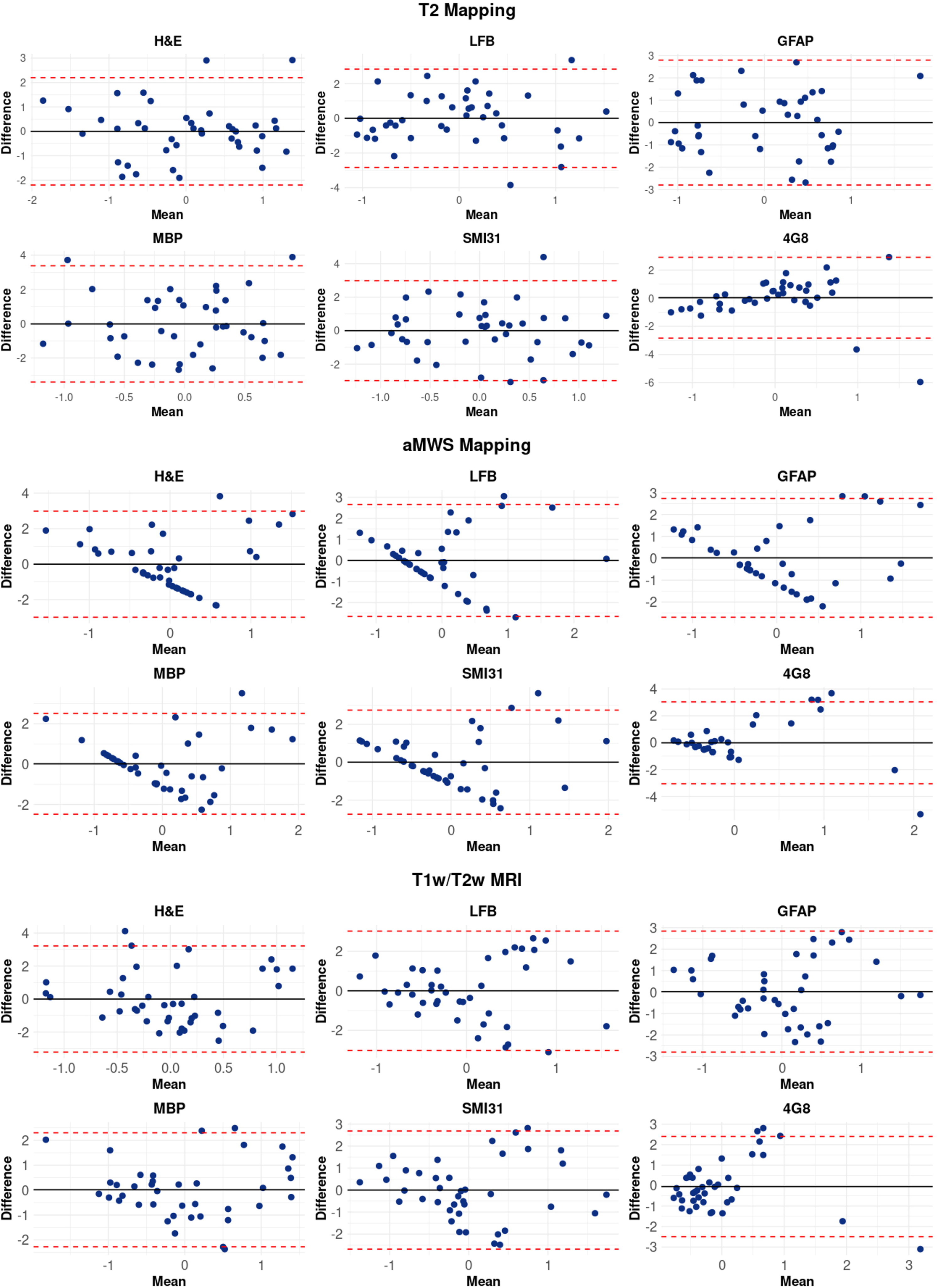
Bland-Altman plots associated with the results in Figure 3.

## 3. Results

The average percent area stained and the corresponding average MRI signal values are presented in Figure 3. These results indicate that WMH exhibit elevated T2 relaxation times, which is expected—greater vacuolation increases interstitial fluid content, leading to prolonged T2 times. Lesions also show reduced aMWS values, consistent with demyelination, suggesting that this method can effectively capture myelin loss. The Bland–Altman plots in Figure 4 demonstrate good agreement between the T2 maps and T1w/T2w MRI, as well as their corresponding histopathology measures. The aMWS maps display a slight negative linear trend, indicating that at higher values, aMWS tends to overestimate the myelin content compared to the histological stains. At lower values, it tends to underestimate it.

Also indicated in Figure 3, T2 times are significantly and positively correlated with vacuolation measured by H&E (p < 0.01, r = 0.46). The aMWS was negatively correlated with vacuolation, with a p-values of 0.01 and r value of -0.41. On the other hand, T2 times were significantly negatively correlated with MBP staining (p < 0.01, r = -0.55), while aMWS and T1w/T2w MRI showed significant positive correlations with MBP (p < 0.01 and p = 0.02, r = 0.46 and r = 0.38, respectively).

The results obtained for the different brain regions are displayed in Figure 5. Notably, decreased MBP percent area and H&E-measured vacuolation align with the qMRI findings. H&E staining reveals lower vacuolation in NAT compared to lesion areas, while MBP staining indicates a higher myelin content in NAT. The NAT exhibits lower T2 values, reflecting preserved tissue integrity. Additionally, NAT shows a higher aMWS signal, further supporting the observed myelin loss within WMH. The T1w/T2w signal is also elevated in NAT, consistent with expected trends.

**Figure 5:**
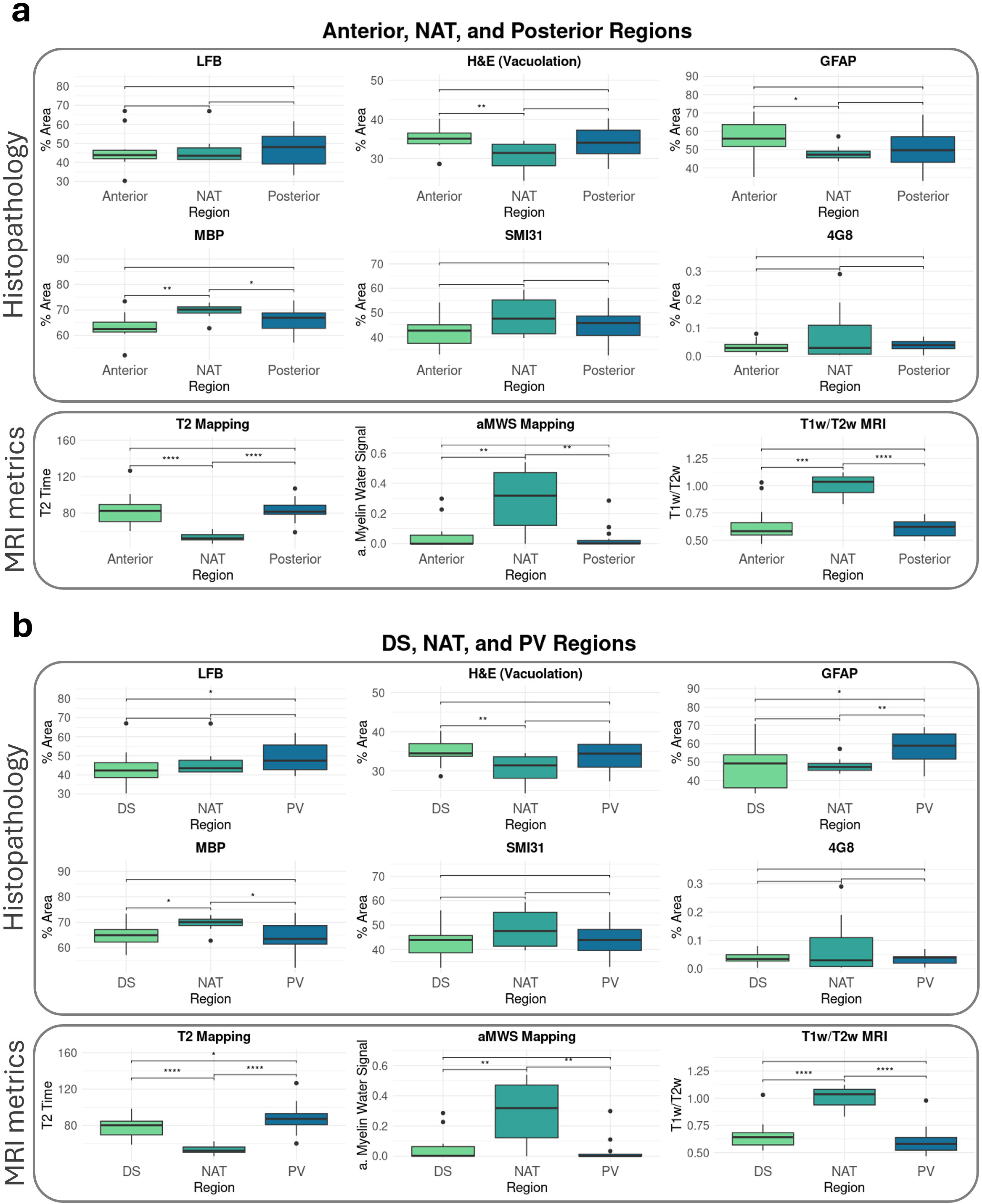
Comparison of the histopathology features and qMRI measures measured in the different brain regions. The anterior and posterior include both periventricular (PV) and deep subcortical (DS) areas (a), while the PV and DS groups include the anterior and posterior regions (b). WMH regions are compared to the normal-appearing tissue (NAT). Asterisks indicate statistically significant differences between regions according to the pairwise two-tailed Wilcoxon rank-sum test. Significance levels are represented as follows: pLJ<LJ0.05 (*), pLJ<LJ0.01 (**), pLJ<LJ0.001 (***), and pLJ<LJ0.0001 (****). The interpretation of each stain’s results in WMH regions compared to NAT is summarized in Table 2. Higher values of LFB, MBP, and SMI31 indicate better white matter integrity. Higher values of H&E, GFAP, and 4G8 indicate worse white matter integrity.

## 4. Discussion

This study provides critical histopathological validation for qMRI metrics in characterizing WMH in AD. By directly aligning MRI-derived metrics with corresponding postmortem histological measures, our findings demonstrate strong associations of both T2 mapping and aMWS mapping with histologically confirmed myelin loss and tissue vacuolation. The Bland-Altman analyses further underscore the reliability of T2 mapping, which displayed superior agreement with histopathology compared to aMWS mapping, the latter showing tendencies toward systematic bias. In comparison, T1w/T2w MRI exhibited weaker and less consistent associations with underlying histopathology, suggesting limited utility in accurately reflecting WMH microstructure. These interpretations must consider the effects of formalin fixation, which biases derived NMR parameters, most notably decreasing T1 and T2 relaxation times and increasing myelin water fraction values (Seifert, et al. 2019, Shatil, et al. 2018). Therefore, further validation on fresh, unfixed samples is still required. Our results collectively form a basis for such future investigation providing evidence of the potential of specific qMRI techniques, particularly T2 mapping and aMWS mapping, as robust, non-invasive biomarkers suitable for clinical application in monitoring white matter pathology and disease progression in neurodegenerative disorders.

### 4.1 T1w/T2w MRI

Both T1-weighted and T2-weighted scans are sensitive to the presence of myelin, but in opposite directions (Bock, et al. 2009). Dividing the T1-weighted and T2-weighted scans increases the contrast-to-noise ratio between myelinated and non-myelinated areas, mathematically canceling the intensity bias from the coils without requiring more complex bias-correcting methods (Glasser and Van Essen 2011). Because of these properties, T1w/T2w MRI has been proposed as a proxy for myelin content. However, prior studies (Arshad, Stanley and Raz 2017, Uddin, Figley and Solar, et al. 2019, Uddin, Figley and Marrie, et al. 2018)—and our findings—suggest a lack of accuracy in the myelin density estimates (Sandrone, et al. 2023). Furthermore, it was established that this ratio is still influenced by scanner differences (Sandrone, et al. 2023). In our study, T1w/T2w ratios showed weaker correlations with histological markers of myelin (H&E and MBP) compared to aMWS and quantitative T2, further questioning its utility as a direct measure of myelin integrity.

### 4.2 Validation of qMRI with Histopathology

Our study is among the first to validate qMRI metrics through direct comparison with values derived from high-resolution histopathological imaging. This approach establishes a crucial link between in vivo MRI signals (Xu, et al. 2025) and the underlying tissue microstructure and pathology, thereby enhancing the interpretability of imaging biomarkers. By employing automated quantification methods for histological staining and systematically assessing their associations with imaging data, our analysis improved robustness and reproducibility.

We observed significant correlations between all three MRI-derived metrics —aMWS mapping, T2 mapping, and T1w/T2w MRI —and histological markers indicative of myelin integrity (MBP and H&E). Notably, these metrics did not show significant correlations with axonal (SMI31) or glial (GFAP) markers. Among the qMRI metrics evaluated, aMWS consistently exhibited the strongest overall association with MBP and H&E staining. However, quantitative T2 mapping slightly outperformed aMWS mapping with respect to MBP correlations in specific regions. The Bland-Altman analyses further underscore the reliability of T2 mapping, revealing superior agreement with histopathology as indicated by a more random distribution around the line of identity. In contrast, aMWS mapping demonstrated tendencies toward systematic bias, suggesting potential limitations in its current implementation. This variability in performance may reflect constraints in earlier versions of our imaging protocol, particularly the limited number of echo times, reducing sensitivity to the short T2 component corresponding to myelin water. Recent protocol enhancements incorporating increased echo sampling are expected to mitigate this limitation, improving the ability to differentiate myelin-bound water from intra- and extracellular compartments in future studies. Our results are consistent with prior studies demonstrating that aMWS mapping correlates strongly with histological myelin content in both 3T and 7T MRI (Xu, et al. 2025).

### 4.3 Histopathological Basis of WMH

Comparisons between WMH and normal-appearing tissue revealed significantly reduced MBP staining and increased vacuolation (indicated by decreased H&E staining) within lesions, consistent with substantial myelin loss and compromised structural integrity. Concurrently, a pronounced increase in GFAP staining was observed in WMH areas, reflecting enhanced glial reactivity. In contrast, axonal markers (SMI31) showed only modest differences between WMH and NAT. Collectively, these patterns suggest that the MRI-visible appearance of WMH predominantly results from demyelination more than axonal degeneration, likely indicating chronic tissue injury and impaired remyelination capacity (Mason, et al. 2001). Consequently, MRI metrics closely aligned with MBP and H&E staining are expected to optimally reflect these underlying pathological changes. Indeed, among the MRI measures evaluated, aMWS mapping demonstrated strong associations with these histological markers, highlighting its sensitivity to white matter pathology and its potential as a reliable imaging biomarker for characterizing WMH in AD. However, the method may have tendencies toward systematic bias, potentially overestimating or underestimating underlying pathological changes.

### 4.4 Regional Differences in WMH Pathology

When comparing periventricular and deep subcortical WMH, no significant differences were observed in MBP or SMI31 staining, suggesting comparable levels of myelination and axonal density between the regions. However, GFAP levels were significantly elevated in periventricular lesions, consistent with prior reports of increased astrocytic and microglial activation in these areas (Simpson, et al. 2007, Murray, et al. 2012). LFB staining also revealed denser myelination in periventricular regions. Although the etiology of these regional differences is beyond the scope of this study, vascular factors, such as those described by Fernando et al. (2006), may contribute to these patterns.

These findings align with epidemiological studies showing that periventricular and subcortical WMH are associated with different clinical outcomes, particularly in AD and vascular dementia (VaD) (Habes, et al. 2018). Our histopathological evidence strengthens the hypothesis that distinct biological mechanisms may contribute to region-specific WMH patterns. This underscores the value of integrating tissue-level validation with imaging data to better interpret the clinical relevance of WMH in neurodegenerative disease.

WMH are often multifactorial in origin. As shown in the postmortem study by McAleese et al. (2021), cerebrovascular pathology frequently coexists with AD, complicating the interpretation of white matter changes on MRI (McAleese, Miah, et al. 2021). Our results highlight the importance of using specific qMRI metrics such as aMWS and T2 mapping to disentangle overlapping pathological processes. By capturing myelin integrity more directly, aMWS mapping may help distinguish between neurodegenerative and vascular contributions to WMH.

## 5. Conclusion

This study provides one of the first direct validations of qMRI measures against histopathology in AD, demonstrating that aMWS mapping and T2 mapping reliably reflect myelin loss and tissue vacuolation within WMH. Compared to T1w/T2w ratios, which showed weaker associations with histological changes, aMWS mapping correlated strongly with myelin content as measured by MBP and H&E staining. According to these results, T2 and aMWS mapping are reliable, non-invasive imaging biomarkers of lesions in the white matter. Additionally, we found that periventricular and subcortical WMH had distinctive gliosis patterns, with periventricular regions showing higher GFAP staining, which may indicate pathophysiological mechanisms specific to that region. By integrating automated histological quantification with matched postmortem MRI, our results underscore the potential of qMRI for capturing demyelination and gliosis, providing a crucial link between imaging and the underlying tissue changes in aging and neurodegenerative disease.

## CRediT Authorship Contribution Statement

**Mariam Mojtabai:** Conceptualization, Formal analysis, Methodology, Writing – original draft, Writing – review & editing. **Rajeswar Kumar:** Writing – original draft, Writing – review & editing. **Nicolas Honnorat:** Conceptualization, Formal analysis, Methodology, Writing – original draft, Writing – review & editing. **Karl Li:** Conceptualization, Formal analysis, Methodology, Writing – original draft, Writing – review & editing. **David H. Wang:** Formal analysis, Investigation, Writing – original draft, Writing – review & editing. **Jinqi Li:** Investigation. **Ray Lee:** Investigation. **Timothy E. Richardson:** Writing – review & editing. **José E. Cavazos:** Writing – review & editing. **Mustapha Bouhrara:** Writing – review & editing. **Susan Resnick:** Writing – review & editing. **Jon B. Toledo:** Writing – review & editing. **Susan Heckbert:** Writing – review & editing. **Margaret E. Flanagan:** Writing – review & editing. **Kevin F. Bieniek:** Formal Analysis, Supervision, Writing – review & editing. **Jamie M. Walker:** Writing – review & editing. **Sudha Seshadri:** Project administration, Resources, Writing – review & editing. **Mohamad Habes:** Conceptualization, Formal analysis, Funding acquisition, Methodology, Project administration, Resources, Supervision, Writing – original draft, Writing – review & editing.

## Acknowledgments

We would like to thank the many generous donors who have graciously donated their brains to our Brain Bank, making this possible. We would also like to thank everyone at the Biggs Institute for helping with collecting all the data necessary to build this repository.

## Data Availability

The corresponding author can be contacted to discuss data sharing.

